# Benign biopsy effect on artificial intelligence cancer detector in screening mammography – a retrospective study

**DOI:** 10.1101/2022.12.24.22283707

**Authors:** Athanasios Zouzos, Alexandra Molovanovic, Karin Dembrower, Fredrik Strand

**Affiliations:** Department of Radiology, Karolinska University Hospital, Stockholm, Sweden; Department of Oncology and Pathology, Karolinska Institute, Stockholm, Sweden; Department of Radiology, S:t Görans Hospital, Stockholm, Sweden

## Abstract

Artificial intelligence (AI) cancer detectors are showing promising results and may soon be used for clinical breast cancer screening in radiology departments. Validation of such programs in different settings is mandatory.

This is a retrospective study after the application of a commercial AI cancer detector program to a cancer-enriched mammography screening dataset where 10 889 women with a median age for the study population of 56 years and a range between 40 and 74 years old where included. The program is intended for 2D mammography and it generate a prediction score for tumor presence with a decimal number between 0 and 1, where 1 represented the highest level of suspicion. The AI score median and interquartile range for women who were healthy, who had a benign biopsy, and who were diagnosed with breast cancer, was calculated. For women with a previous benign biopsy, the time between mammogram and the biopsy was stratified. The effect of increasing age was examined using stratified analysis and linear regression modelling. The proportion of healthy women above the AI abnormality threshold (0.4) defined at our hospital was 3.5%, 11% and 84% for healthy women without a benign biopsy, healthy women with a benign biopsy, and for women with breast cancer, respectively (p<0.001) and the AI score correlated positively with increasing age of the women in the cancer group (p<0.05). While 16% of the women who had a benign biopsy after mammography had an AI score above the cut-off point, there were 33% of those when they had their biopsy before the mammography. AI cancer detection systems can reliable be used in a screening setting. Adding information about possible prior benign biopsy and other symptoms can probably improve accuracy and subsequently reduce the workload for radiologists.

## Introduction

Breast cancer is the most common cancer among women worldwide. It ranks 5th as a cause of cancer deaths because of its relatively favorable prognosis but in the last 20 years the average annual increase in breast cancer incidence rate has been 1.4% [1-3].

Well implemented screening programs have been clearly proven to reduce the mortality rate for breast cancer [4-6] and prospective studies have shown that results improve when radiologists combine mammography reading with an AI algorithm [7-9]. Furthermore, reducing reading time with the assistance of AI algorithms is possible [10, 11]. AI algorithms can be highly accurate for reading mammograms and some systems are now on a comparable level with average breast radiologists at detecting breast cancer on screening mammography [12].

In addition to the well-known risk factors of age, family history and hormonal history, there are also studies showing that benign breast disease increases breast cancer risk [13, 14]. A study that analyzed risk factors for breast cancer found that having had any prior breast procedure was associated with an increased risk of breast cancer [15]. Another study showed that women found to have false-positive mammography were more likely to develop interval cancer or cancer at the second screening compared to women not recalled [16]. Also in Gail’s risk model, prior biopsy is defined as a risk factor, and the risk is higher for women under 50 years of age [17].

In light of biopsies as a breast cancer risk factor and the increasing use of AI for screening mammography, it is important to understand if having a benign biopsy affects the AI cancer score and the ability of AI to make an accurate assessment of the screening mammogram.

## Results

Initially there were 11 303 women in this retrospective case-control study. Of these 414 women were excluded. The exclusion criteria were; no mammography in conjunction with a cancer diagnosis, implants and cancer more than 12 months after mammography. The cancer group consisted in total of 917 women, the benign biopsy-group of 234 women and in the group with no cancer or biopsy there were 9 738 women, see Fig 1.

**Fig 1.**
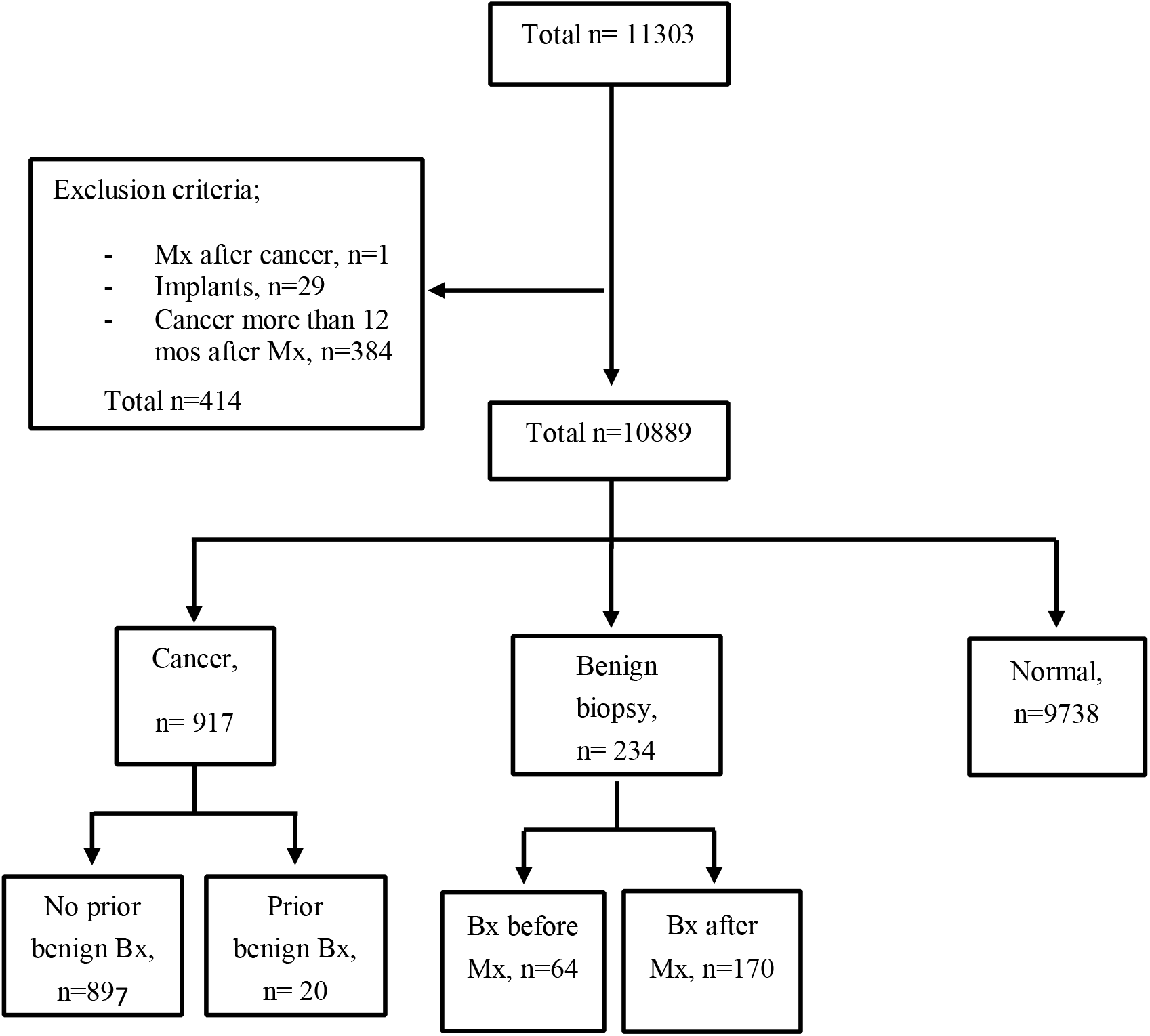
Study population with exclusion criteria and subgroups. Bx: benign biopsy; Mx: mammography.

Of the remaining 10 889 women, 8 307 had complete information on radiologist assessments (when performing data collection, we received radiologist assessments only until Dec 31, 2015) which included selections as potentially pathological by one or both radiologists and a final recall decision after consensus discussion. From those 8 307 women there where 724 women in the cancer group, 212 in the benign biopsy-group and 7 371 in the normal mammographygroup.

There was a significant difference (p<0.001) in AI score between the cancer-group, the benign biopsy-group and the normal mammography-group (Table 1).

**Table 1.**
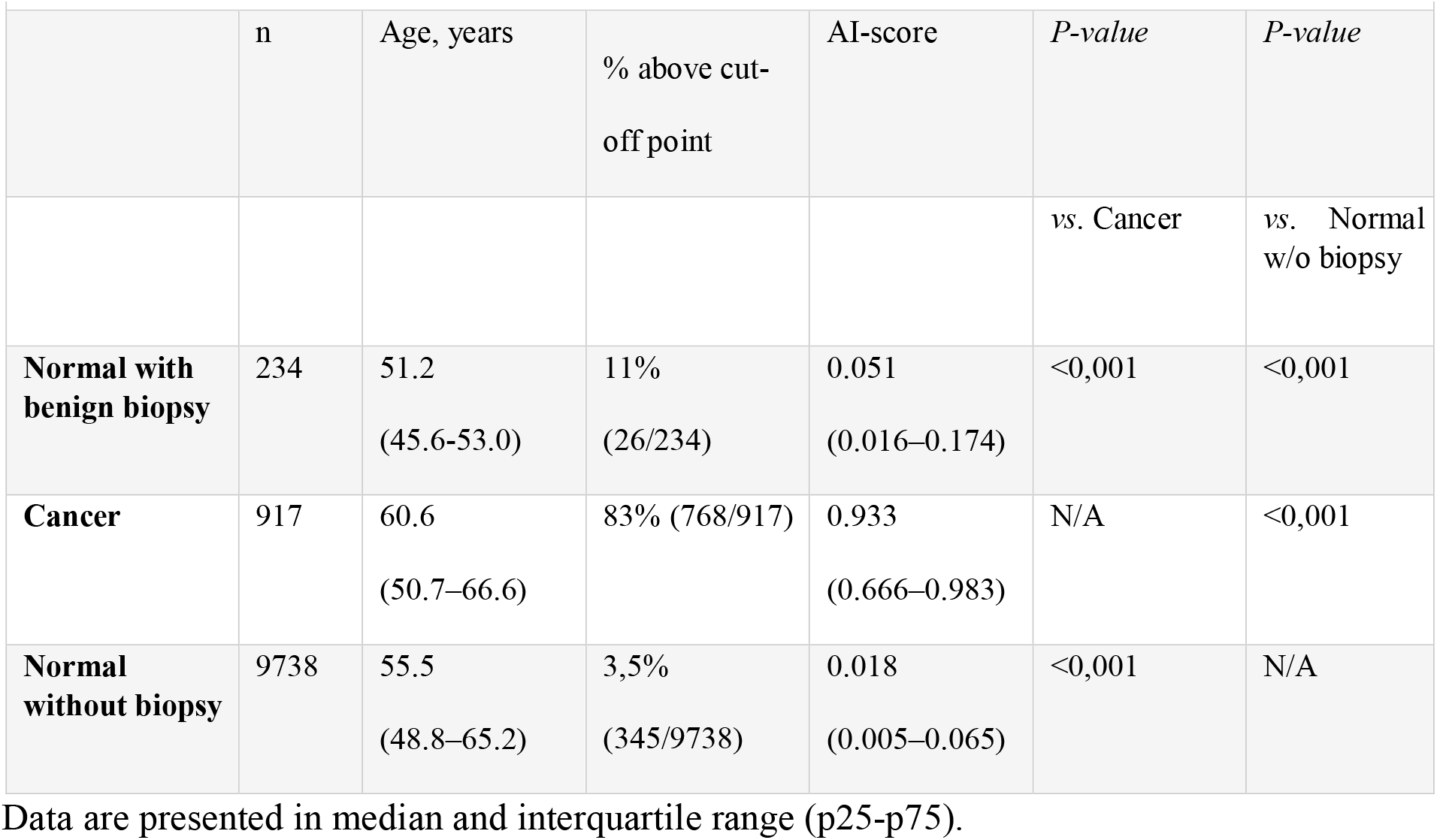
Study population – Sub groups.

|  | n | Age, years | % above cut-off point | AI-score | *P-value* | *P-value* |
| --- | --- | --- | --- | --- | --- | --- |
|  |  |  |  |  | vs. Cancer | vs. Normal w/o biopsy |
| **Normal with benign biopsy** | 234 | 51.2<br>(45.6–53.0) | 11%<br>(26/234) | 0.051<br>(0.016–0.174) | <0,001 | <0,001 |
| **Cancer** | 917 | 60.6<br>(50.7–66.6) | 83% (768/917) | 0.933<br>(0.666–0.983) | N/A | <0,001 |
| **Normal without biopsy** | 9738 | 55.5<br>(48.8–65.2) | 3,5%<br>(345/9738) | 0.018<br>(0.005–0.065) | <0,001 | N/A |

Data are presented in median and interquartile range (p25–p75).

The percentage above the cut-off point, for AI assessments, in the group with normal mammography was 3.5%, and 83% in the cancer-group. In the benign biopsy-group 11% of the AI assessments were above the cut-off point. The distribution of AI scores for women diagnosed with breast cancer is shown in Fig 2, for healthy women with benign biopsy in Fig 3, and for healthy women without benign biopsy who remained healthy in Fig 4.

**Fig 2.**
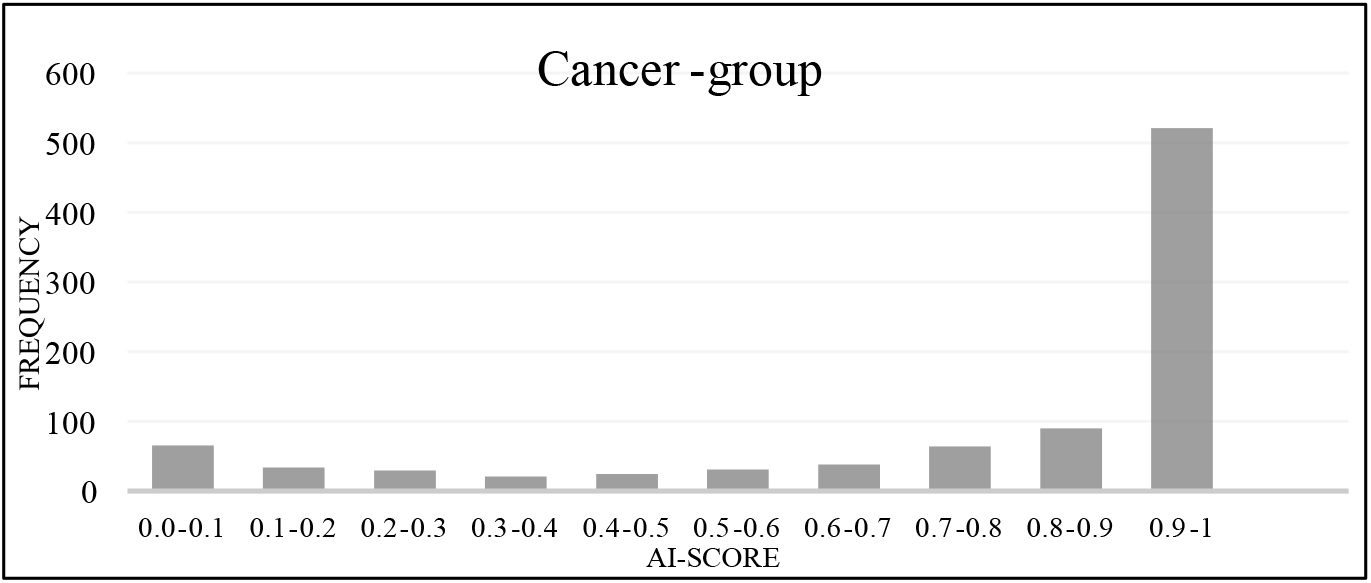
N=917. Median 0.933, p25-p75 (0.666-0.983). Cut-off point 0.40

**Fig 3.**
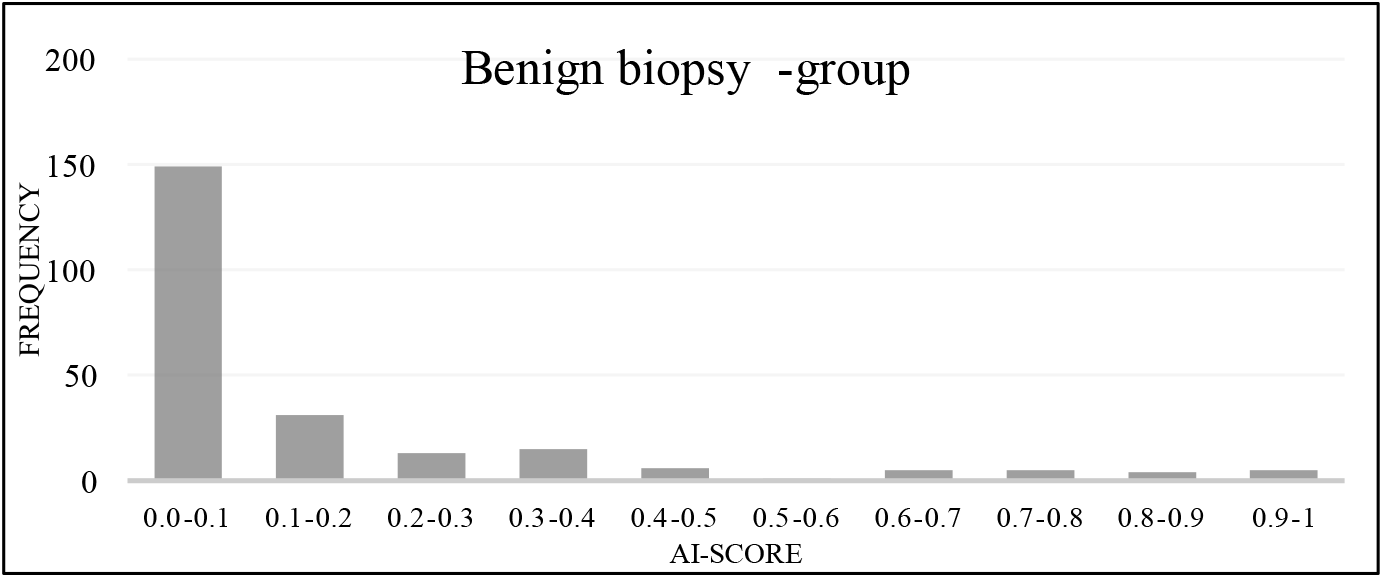
N=234. Median 0.051, p25-p75 (0.016-0.174). Cut-off point 0.40

**Fig 4.**
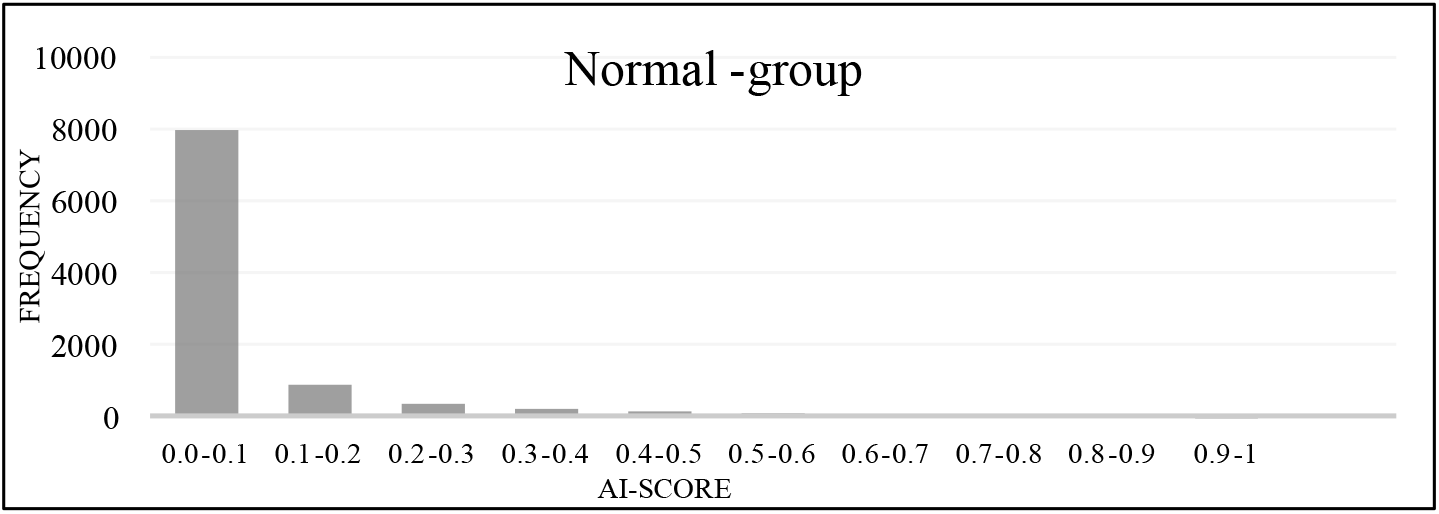
N=9738. Median=0.018. p25-p75 (0.005-0.065). Cut-off point 0.40

In Table 2, we present how the AI score is associated with the age of the woman. There was a significant increase of the AI score in relation to age category in the cancer group (*p*<0.05). There was no significant increase in the AI score in relation to age in the group with normal mammography or in the group with benign biopsies. The median age for the study population was 56 years and the median AI score was 0.023. The median age for the cancer group was 61 years and the median AI score was 0.933. The median age for the group with previous benign biopsies was 49 years and the median AI score 0.051. The median AI score for healthy women was 0.018 and the median age for that group was 59 years.

**Table 2.** AI-score, age order

| Age (years) | Normal mammography (No bx, no cancer) |  | Benign biopsy-group (No cancer) |  |  |  |  |  | Cancer-group |  |  |  |  |  |
| --- | --- | --- | --- | --- | --- | --- | --- | --- | --- | --- | --- | --- | --- | --- |
|  |  |  | All |  | Bx before Mx |  | Bx after Mx |  | All |  | Prior benign Bx |  | No benign Bx |  |
| **All** | 9738 | 0.018<br>(0.005–0.065) | 234 | 0.051<br>(0.016–0.174) | 64 | 0.069<br>(0.018–0.179) | 170 | 0.047<br>(0.016–0.205) | 917 | 0.933<br>(0.667–0.983) | 20 | 0.924<br>(0.747–0.986) | 897 | 0.934<br>(0.666–0.983) |
| **40–49** | 3152 | 0.021<br>(0.006–0.068) | 144 | 0.048<br>(0.016–0.155) | 43 | 0.074<br>(0.019–0.142) | 101 | 0.042<br>(0.012–0.176) | 213 | 0.861<br>(0.272–0.974) | 9 | 0.861<br>(0.286–0.974) | 204 | 0.864<br>(0.264–0.975) |
| **50–59** | 2834 | 0.015<br>(0.005–0.058) | 68 | 0.051<br>(0.017–0.260) | 16 | 0.039<br>(0.014–0.113) | 52 | 0.051<br>(0.017–0.305) | 228 | 0.948<br>(0.715–0.985) | 3 | 0.911<br>(0.891–0.950) | 225 | 0.949<br>(0.709–0.985) |
| **60–69** | 2577 | 0.018<br>(0.005–0.064) | 22 | 0.078<br>(0.012–0.174) | 5 | 0.173<br>(0.090–0.449) | 17 | 0.047<br>(0.011–0.144) | 379 | 0.935<br>(0.723–0.984) | 8 | 0.945<br>(0.887–0.991) | 371 | 0.935<br>(0.720–0.984) |
| **70+** | 1175 | 0.020<br>(0.006–0.077) | 0 |  | 0 |  | 0 |  | 97 | 0.958<br>(0.820–0.985) | 0 |  | 97 | 0.958<br>(0.820–0.985) |

Data are presented in median with interquartile range (p25–p75). Bx= benign biopsy,
Mx=mammography

The benign biopsy-group was stratified by time between biopsy and mammography into three categories: 0-6 months, 6-24 months and more than 24 months. There was no significant difference between the time-stratified categories (Table 3). In Table 3, we describe the AI score related to the time between biopsy and mammography. Within 6 months after mammography, 104 of 234 had had benign biopsy. The proportion of women with AI scores above the threshold, was, 16% for those with a benign biopsy within 6 months from mammography, and 33% for those with a benign biopsy 6 months before mammography.

**Table 3.** Study population - Benign biopsies.

|  | n | Age, years | % above the cut-off point | AI-score | *P-value* |
| --- | --- | --- | --- | --- | --- |
| **Benign biopsy before mammography** |  |  |  |  |  |
| **0-6 months** | 9 | 49.1<br>(44.5-54.9) | 33%<br>(3/9) | 0.150<br>(0.099-0.449) | ref |
| **6-24 months** | 39 | 48.1<br>(46.7-52.7) | 5.1% (2/39) | 0.062<br>(0.016-0.150) | 0.123 |
| **>24 months** | 16 | 41.0<br>(40.3-48.4) | 0% | 0.025<br>(0.015-0.099) | 0.055 |
| **Benign biopsy after mammography** |  |  |  |  |  |
| **0-6 months** | 104 | 48.3<br>(44.4-52.5) | 16%<br>(17/104) | 0.065<br>(0.017-0.274) | ref |
| **6-24 months** | 44 | 49.0<br>(45.6-55.3) | 4.5% (2/44) | 0.037<br>(0.008-0.109) | 0.362 |
| **>24 months** | 22 | 51.2<br>(45.6-53.0) | 9%<br>(2/22) | 0.031<br>(0.017-0.165) | 0.378 |

Data are presented in median and interquartile range (p25-p75).

In the group with benign biopsy after mammography the abnormal assessment by AI, above cut-off point, was 15% while the radiologists had a recall rate up to 57% in this group (Table 4).

**Table 4.** Recall rate and abnormal assessments.

|  | Radiologist 1 | Radiologist 2 | Consensus | AI-% above cut-off point |
| --- | --- | --- | --- | --- |
| **Total** | 10%<br>(831/8307) | 10%<br>(827/8307) | 9.2%<br>767/8307) | 11% (880/8307) |
| **Normal and**<br>**benign biopsy** | 3.6%<br>(274/7583) | 3.1%<br>(234/7583) | 2%<br>(154/7583) | 3.8%<br>(290/7583) |
| **Normal** | 2.8%<br>203/7371) | 2.2%<br>(162/7371) | 1%<br>(73/7371) | 3.6%<br>(265/7371) |
| **Benign biopsy** | 33%<br>(71/212) | 34%<br>(72/212) | 38%<br>(81/212) | 12%<br>(25/212) |
| **- Biopsy before Mx** | 8.5% (5/59) | 6.8% (4/59) | 6.8% (4/59) | 8.5% (5/59) |
| **- Biopsy after Mx** | 49%<br>(66/135) | 50%<br>(68/135) | 57%<br>(77/135) | 15%<br>(20/135) |
| **Cancer** | 77%<br>(557/724) | 82%<br>(593/724) | 85%<br>(613/724) | 81% (590/724) |
| **With benign biopsy** | 75%<br>(12/16) | 88%<br>(14/16) | 88%<br>(14/16) | 81%<br>(13/16) |
| **W/o benign biopsy** | 77%<br>(545/708) | 82%<br>(579/708) | 85%<br>(599/708) | 82% (577/708) |

Recall rate by radiologist and abnormal assessments by AI. Data are presented in percentage and fraction. Mx: mammography.

The radiologists had a recall rate of 2% and the rate for abnormal assessments by AI was 3.8% in the group with normal mammograms and with benign biopsy (Table 4). For the group with only normal mammograms the recall rates were 1% and 3.6% respectively. Radiologists and the AI program had similar rates of recall for the total study population as a whole.

The following two screening mammograms have been assessed by radiologists and the AI cancer detection program, see Figs 5 and 6. These examples illustrate concordant and discordant assessments.

**Fig 5.**
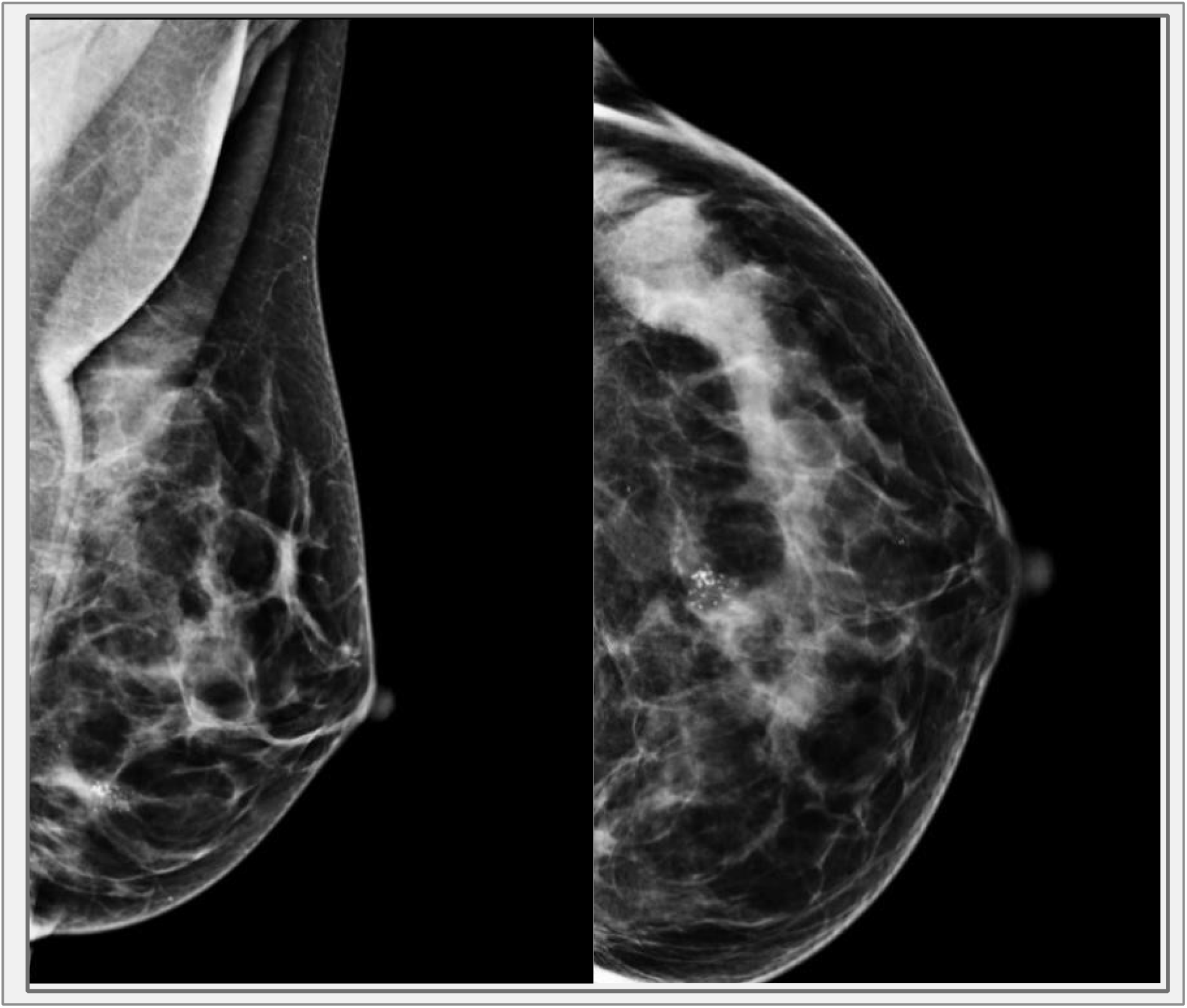
Woman in her 50s, selected by radiologists for potential pathology in the left breast. High AI-score. The biopsy result showed hyperplastic breast epithelial cells that could represent a degenerated fibroadenoma.

**Fig 6.**
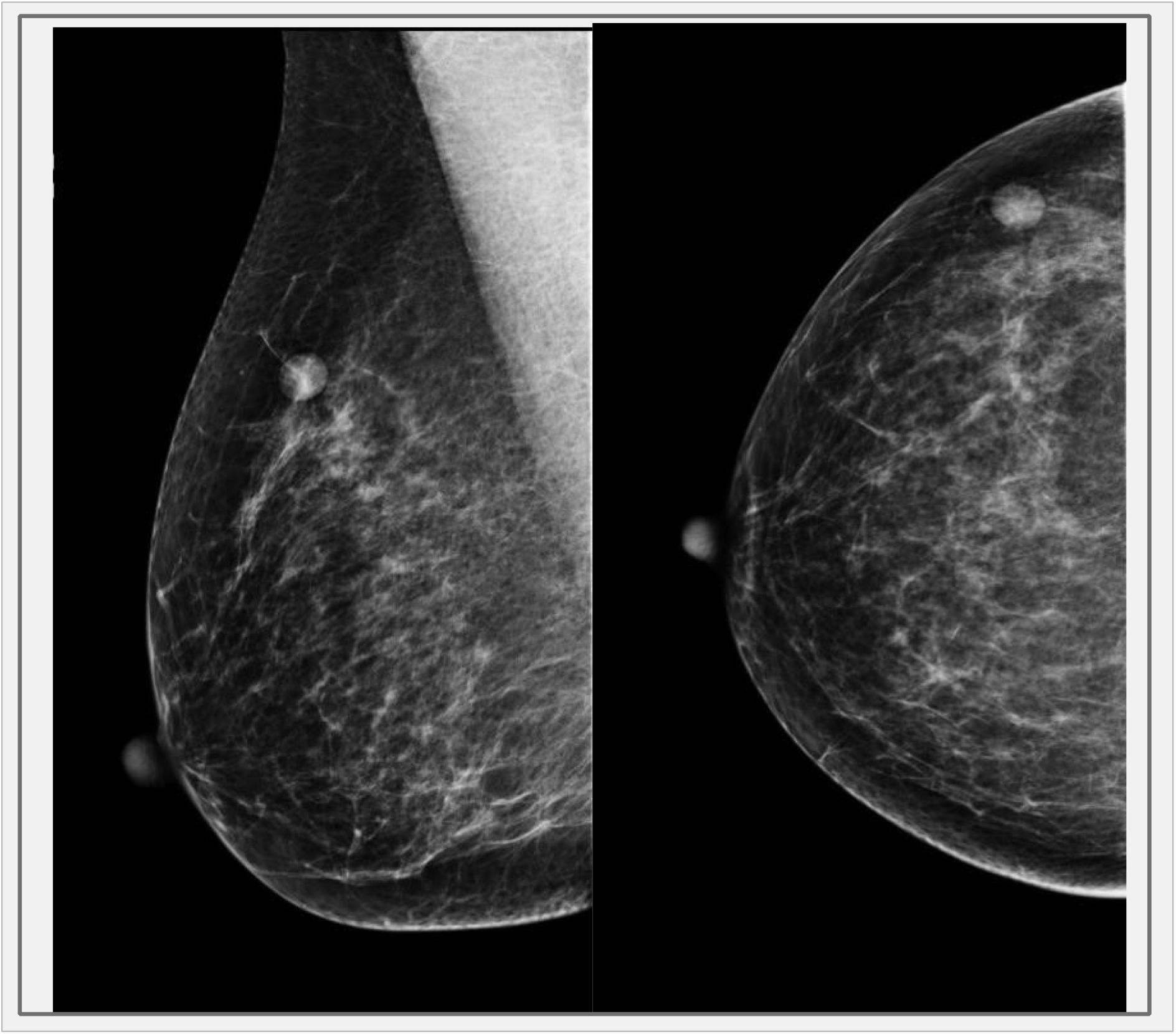
Woman in her 50s, selected by radiologists for potential pathology in the right breast. Low AI-score. The biopsy showed lymph node.

## Material and method

### Study population

This project is a retrospective study based on a case-control subset from the dataset Cohort of Screen-Aged Women (CSAW). CSAW is a complete population-based cohort of women 40 to 74 years of age invited to screening in the Stockholm region, Sweden, between 2008 and 2015 [18]. The exclusion criteria in CSAW were: prior history of breast cancer, diagnosis outside the screening range and incomplete mammography examinations. From this large CSAW-cohort a case-control subset was separately defined to contain all women from Karolinska University Hospital, Stockholm, who were diagnosed with breast cancer (n=1303), at screening or clinically during the interval before the next planned screening, and 10 000 randomly selected healthy controls [18]. The purpose of the case-control subset is to make evaluation more efficient by not having to process an unnecessary amount of healthy controls while preserving the representability of the CSAW screening cohort in which it is nested. Additional exclusion criteria for the current study were: implants and cancer diagnosis more than 12 months after mammography. The study population was divided into three groups based on their status: cancer, benign biopsy, and normal.

The cancer-group was defined as “biopsy verified breast cancer within 12 months from mammography”. For those women, the most recent screening mammography prior to diagnosis was selected for analysis.

The benign biopsy-group was defined as “benign biopsy without ever having breast cancer”. The screening mammography closest in time to the biopsy, before or after was chosen. The group was stratified by time between biopsy and mammography, both from biopsy to mammography and from mammography to biopsy.

The normal-group had had neither breast cancer nor prior benign biopsy. The most recent screening mammography was selected for this group. Women in the screening program with lesions that were previously deemed benign and were not selected for further diagnostic work, were also included in this group. Only biopsy verified lesions were included in the benign biopsy-group.

### Mammography assessments

Screening mammograms were prospectively assessed independently by two radiologists working at a certified screening center, with more than five years of screening experience for each of them. The following screening decision data was collected: flagging of abnormal screening by one or both radiologists and the final recall decision after consensus discussion. Screening decisions and clinical outcome data was collected by linking to regional cancer center registers. Those data were collected so to be compared with the assessment results from AI cancer detector.

The AI cancer detector algorithm used for detection of tumor signs was provided by a commercial vendor (Lunit, Seoul, South Korea), version 5.5.0.16. The reason for choosing Lunits algorithm for this study was the results of the retrospective analysis published 2020 [9] where a comparison of three of the most known algorithms showed that best of those was AI1 (Lunit) with a sensitivity and specificity comparable to Breast Cancer Surveillance Consortium benchmarks [19].

The algorithm was originally trained on 170 230 mammograms, from 36 468 women diagnosed with breast cancer and 133 762 healthy controls. The mammograms in the original training were sourced from five institutions: three from South Korea, one from the USA, and one from the UK. The mammograms were acquired on mammography equipment from GE Healthcare, Hologic, and Siemens, and there were both screening and diagnostic mammograms. In the clinical version, the AI cancer-detector software creates visual prompts in the mammograms where the suspicion of tumor is above a certain threshold. In this study, we did not use the images, but instead used the underlying continuous prediction score of the algorithm. The generated prediction score for tumor presence was a decimal number between 0 and 1, where 1 represented the highest level of suspicion. The program assessed both breasts and the highest score among the four images was selected as representing the exam. The cut-off point (0.4) (AI abnormality threshold) defining a screening abnormality was determined in a prior study having the AI system operate at the first radiologists’ specificity level [9].

### Collection of data

The Stockholm-Gotland regional cancer center (RCC) provided personal identification numbers for all women who fulfilled the inclusion criteria for CSAW. The identification numbers were linked to the local breast cancer quality register, “Regional Cancercentrum Stockholm-Gotlands Kvalitetsregister för Bröstcancer” to collect data about breast cancer diagnosis. All diagnoses of breast cancer were biopsy verified. Benign diagnoses were collected from the medical record “Take Care” which is hospitals data-based system that provides detailed health information about every registered patient.

All images were 2D full-field digital mammograms acquired on Hologic mammography equipment. The personal identification numbers were also linked to the radiological image repository to extract all digital mammograms from PACS. The dataset has been stored securely following the requirements of the ethical approval.

### Statistical analyses

The statistical analysis was done per patient and not per lesion. Stata version 14 or later was used for statistical analyses. Wilcoxon rank sum-test and quantile regression were used to examine differences between groups. These statistical tests of differences in median were chosen due to the skewed distribution of AI scores.

### Compliance with ethical standards

The collection and use of the dataset for the purpose of AI has been approved by the Swedish Ethical Review Board 2017-02-08, and the need for informed consent was waived.

The Diary number was 2016/2600-31.

## Discussion

The strength of this study is the large number of cancer cases and that all women were sampled from a screening cohort. A limitation is the relatively small number of benign biopsies which makes it difficult to consider different types of benign lesions. Another limitation is the retrospective setting: since the AI program did not have the opportunity to make recalls and choose women for further diagnostic biopsy it could not affect who got biopsied. All information about benign biopsies was based on radiologists’ decisions. In contrast to radiologists, the AI-program calculates a score for the likelihood of breast cancer based on the image alone and does not consider any information about symptoms given by the woman at screening.

Furthermore, in this study, we did not consider the exact location of the presumed abnormality where the AI showed a high AI score. We did not use the images, but instead we used the underlying predictions score of the algorithm as a per patient analysis. AI scores above the threshold for women having benign biopsy was 16% if the biopsy was done 6 months after the mammography and 33% if the biopsy was done 6 months before the mammography examination. This can cause questions about the probability that AI is affected by alternations on mammography because of the biopsy. Further analysis of the data can be valuable to evaluate whether the lesions that AI showed responded to the actual finding that the patient was recalled for.

Although we used a specific AI cancer-detector software for our study which may reduce the external validity of the study, the mammograms were acquired on mammography equipment from GE Healthcare, Hologic, and Siemens, and there were both screening and diagnostic mammograms which covers a broad amount of systems with diverse imaging characteristics. The data that confirm the external validation of the AI program is the significant difference in AI score between the normal group and the cancer group as well as the association between higher AI score and increasing age, in the cancer-group. Like radiologists, AI detects more cancer in the screening mammograms of older women. No difference was shown between AI assessment and radiologist assessment when selecting for cancer in accordance with results from prior studies.

Another interesting finding is the recall rate for the group with mammography prior to benign biopsy. It was 57% for the radiologist and 15% for the AI program. Those figures are more likely to be explained by the study design where the AI program didn’t have access to the clinical evaluation of the patients while the radiologists selected all the patients with symptoms as in routine praxis almost independently from imaging.

The most intriguing finding this study presents is that even if the proportion of (false positive) AI scores increased from 3.6% to 8.5% with a previous benign biopsy. In other words, there was a significant difference in AI scores between the normal group and the benign biopsy-group despite both groups consisting of healthy women. This is probably an indication that incorporation of information about prior biopsy in AI cancer detector programs would further improve their accuracy. Prospective studies regarding that matter are required.

These results amplify the indications from previous studies, that AI cancer detectors can be reliable enough to incorporate in a clinical screening assessment. If AI is to make more accurate assessments it will probably require clinical information (previous biopsy, symptoms) in addition to simply analyzing mammograms. Based on the observation that only 15% of the women who had a biopsy after mammography had an AI score above the cut-off point, there might be a role for AI in reducing the number of unnecessary biopsies.

## Data Availability

All our data are available according to PLOS Data Policy

## References

1. Wigzell K, Rosen M. Socialstyrelsen - The National Board of Health and Welfare/Centre for Epidemiology - Foreword. Scandinavian journal of public health. 2001:1.

2. Ferlay J, Soerjomataram I, Dikshit R, Eser S, Mathers C, Rebelo M, et al. Cancer incidence and mortality worldwide: Sources, methods and major patterns in GLOBOCAN 2012. International journal of cancer. 2015;136(5):E359–E86. doi: 10.1002/ijc.29210.

3. Sung H, Ferlay J, Siegel RL, Laversanne M, Soerjomataram I, Jemal A, et al. Global Cancer Statistics 2020: GLOBOCAN Estimates of Incidence and Mortality Worldwide for 36 Cancers in 185 Countries. CA Cancer J Clin. 2021;71(3):209–49. Epub 20210204. doi: 10.3322/caac.21660. PubMed PMID: 33538338.

4. Nyström L, Wall S, Rutqvist LE, Lindgren A, Lindqvist M, Rydén S, et al. Breast cancer screening with mammography: overview of Swedish randomised trials. The Lancet (British edition). 1993;341(8851):973–8. doi: 10.1016/0140-6736(93)91067-V.

5. Marmot MG, Altman DG, Cameron DA, Dewar JA, Thompson SG, Wilcox M. The benefits and harms of breast cancer screening: an independent review. Br J Cancer. 2013;108(11):2205–40. Epub 20130606. doi: 10.1038/bjc.2013.177. PubMed PMID: 23744281; PubMed Central PMCID: PMCPMC3693450.

6. Long H, Brooks JM, Harvie M, Maxwell A, French DP. Correction: How do women experience a false-positive test result from breast screening? A systematic review and thematic synthesis of qualitative studies. Br J Cancer. 2021;125(7):1031. Epub 2021/08/01. doi: 10.1038/s41416-021-01503-w. PubMed PMID: 34331024; PubMed Central PMCID: PMCPMC8476612.

7. Roiguez Ruiz A, Krupinski E, Mordang JJ, Schilling K, Heywang-Kobrunner SH, Sechopoulos I, et al. Detection of Breast Cancer with Mammography: Effect of an Artificial Intelligence Support System. Radiology. 2019;290(2):305–14. doi: 10.1148/radiol.2018181371.

8. Schaffter T, Buist DSM, Lee C, Nikulin Y, Ribli D, Guan Y, et al. Evaluation of Combined Artificial Intelligence and Radiologist Assessment to Interpret Screening Mammograms. JAMA network open. 2020;3(3):e200265–e. doi: 10.1001/jamanetworkopen.2020.0265.

9. Salim M, Wåhlin E, Dembrower K, Azavedo E, Foukakis T, Liu Y, et al. External Evaluation of 3 Commercial Artificial Intelligence Algorithms for Independent Assessment of Screening Mammograms. JAMA Oncol. 2020;6(10):1581–8. Epub 2020/08/28. doi: 10.1001/jamaoncol.2020.3321. PubMed PMID: 32852536; PubMed Central PMCID: PMCPMC7453345 outside the submitted work. Dr Eklund reported receiving grants from Swedish Research Council and from the Swedish Cancer Society during the conduct of the study. Dr Strand reported receiving grants from Stockholm City Council during the conduct of the study; and receiving personal fees from Collective Minds Radiology outside the submitted work. No other disclosures were reported.

10. Roiguez Ruiz A, Lang K, Gubern Merida A, Teuwen JJB, Broeders MJM, Gennaro G, et al. Can we reduce the workload of mammographic screening by automatic identification of normal exams with artificial intelligence? A feasibility study. European radiology. 2019;29(9):4825–32. doi: 10.1007/s00330-019-06186-9.

11. Dembrower K, Wåhlin E, Liu Y, Salim M, Smith K, Lindholm P, et al. Effect of artificial intelligence-based triaging of breast cancer screening mammograms on cancer detection and radiologist workload: a retrospective simulation study. The Lancet Digital health. 2020;2(9):e468–e74. doi: 10.1016/S25897500(20)30185-0.

12. Roiguez Ruiz A, Lang K, Gubern-Merida A, Broeders MJM, Gennaro G, Clauser P, et al. Stand-Alone Artificial Intelligence for Breast Cancer Detection in Mammography: Comparison With 101 Radiologists. JNCI : Journal of the National Cancer Institute. 2019;111(9):916–22. doi: 10.1093/jnci/djy222.

13. Dumitrescu RG, Cotarla I. Understanding breast cancer risk - where do we stand in 2005? Journal of cellular and molecular medicine. 2005;9(1):208–21. doi: 10.1111/j.1582-4934.2005.tb00350.x.

14. Hartmann LC, Sellers TA, Frost MH, Lingle WL, Degnim AC, Ghosh K, et al. Benign Breast Disease and the Risk of Breast Cancer. The New England journal of medicine. 2005;353(3):229–37. doi: 10.1056/NEJMoa044383.

15. Barlow WE, White E, Ballard-Barbash R, Vacek PM, Titus-Ernstoff L, Carney PA, et al. Prospective Breast Cancer Risk Prediction Model for Women Undergoing Screening Mammography. JNCI : Journal of the National Cancer Institute. 2006;98(17):1204–14. doi: 10.1093/jnci/djj331.

16. McCann J, Stockton D, Godward S. Impact of false-positive mammography on subsequent screening attendance and risk of cancer. Breast cancer research : BCR. 2002;4(5):R11–R. doi: 10.1186/bcr455.

17. Gail MH, Brinton LA, Byar DP, Corle DK, Green SB, Schairer C, et al. Projecting Individualized Probabilities of Developing Breast Cancer for White Females Who Are Being Examined Annually. JNCI : Journal of the National Cancer Institute. 1989;81(24):1879–86. doi: 10.1093/jnci/81.24.1879.

18. Dembrower K, Lindholm P, Strand F. A Multi-million Mammography Image Dataset and Population-Based Screening Cohort for the Training and Evaluation of Deep Neural Networks-the Cohort of Screen-Aged Women (CSAW). Journal of digital imaging. 2020;33(2):408–13. doi: 10.1007/s10278-019-00278-0.

19. Lehman CD, Arao RF, Sprague BL, Lee JM, Buist DS, Kerlikowske K, et al. National Performance Benchmarks for Modern Screening Digital Mammography: Update from the Breast Cancer Surveillance Consortium. Radiology. 2017;283(1):49–58. Epub 20161205. doi: 10.1148/radiol.2016161174. PubMed PMID: 27918707; PubMed Central PMCID: PMCPMC5375631.

